# Prostate-specific membrane antigen is a biomarker for residual disease following neoadjuvant intense androgen deprivation therapy in prostate cancer

**DOI:** 10.1101/2021.10.28.21265614

**Authors:** John R. Bright, Rosina T. Lis, Anson T. Ku, Nicholas T. Terrigino, Shana Y. Trostel, Nicole V. Carrabba, Stephanie A. Harmon, Baris Turkbey, Scott Wilkinson, Adam G. Sowalsky

## Abstract

Neoadjuvant intense androgen deprivation therapy can exert a wide range of histologic responses, which in turn are reflected in the final prostatectomy specimen. Accurate identification and measurement of residual tumor volumes are critical for tracking and stratifying patient outcomes. The goal of this current study was to evaluate the ability of antibodies against prostate-specific membrane antigen (PSMA) to detect residual tumor in a cohort of 35 patients treated with androgen deprivation therapy plus enzalutamide for six months prior to radical prostatectomy. Residual carcinoma was detected in 31 patients, and PSMA reacted positively with tumor in all cases. PSMA staining was 95.5% sensitive for tumor, with approximately 81.6% of benign regions showing no reactivity. By contrast, PSMA positively reacted with 72.2% of benign regions in a control cohort of 37 untreated cases, resulting in 27.8% specificity for tumor. PSMA further identified highly dedifferentiated prostate carcinomas including tumors with evidence of neuroendocrine differentiation. We propose that anti-PSMA immunostaining be a standardized marker for identifying residual cancer in the setting of neoadjuvant intense androgen deprivation therapy.

## INTRODUCTION

Strategies for the definitive treatment of intermediate and high-risk localized prostate cancer includes surgery or radiotherapy. However, with rates of biochemical recurrence of 15-30%, the use of neoadjuvant therapies prior to surgery involving hormonal, chemotherapeutic, or precision agents may improve outcome in the subset of unfavorable-risk patients harboring micrometastatic disease at the time of diagnosis [1-5]. Intense androgen deprivation therapy (iADT), which couples an Androgen Receptor (AR) pathway-targeting drug with conventional luteinizing hormone releasing hormone (LHRH) agonists has shown strong efficacy in metastatic, newly-diagnosed hormone-sensitive prostate cancers; iADT is amongst the most frequently used strategies in the locally advanced stage as a neoadjuvant systemic therapy [3, 6].

Although data from recent clinical trials have not yet matured to allow for determining overall survival benefits, a subset of patients receiving neoadjuvant iADT demonstrate significant delays in biochemical recurrence (BCR) based on when they would otherwise be expected to recur relative to historical controls [7, 8]. Indeed, exceptionally responding patients harboring minimal residual disease (or complete pathologic responses) after neoadjuvant therapy, which has been defined as < 0.25-0.5 cm in the largest cross-sectional dimension, or 0.05-0.25cm^3^ by volume, have 3-year BCR-free rates greater than 95%, and thus residual cancer volumes have become primary outcomes of new clinical studies [8]. As the definition of an “incomplete responder” or “nonresponder” (with greater than minimal residual disease) is based on the accurate quantification of remaining cancer cells from a meticulous examination of prostatectomy tissue after surgery, various immunostains are recommended in conjunction with H&E staining [9].

In this study, we hypothesized that immunohistochemistry (IHC) against PSMA would be a sensitive and specific marker for detecting and measuring residual prostate tumor following neoadjuvant iADT. We show that prostate tumors treated with six months of AR-targeted neoadjuvant therapy retain high levels of PSMA expression while benign glands do not. As a result, IHC against PSMA is nearly three-times more specific for tumor cells after treatment, compared to a series of untreated controls. PSMA expression was also retained in cases that had undergone three distinct forms of neuroendocrine differentiation. We thus propose that anti-PSMA IHC be considered as a standard histologic marker for measuring residual cancer volumes in the setting of neoadjuvant iADT.

## MATERIALS AND METHODS

### Biospecimen procurement

The collection and analysis of tissue and demographic data from patients with high-risk localized prostate cancer treated with ADT plus enzalutamide prior to surgery was approved by the National Institutes of Health Institutional Review Board (protocol 15-c-0124). The collection and analysis of tissue and demographic data from patients with localized prostate cancer treated only by surgery was approved by the institutional review boards of Beth Israel Deaconess Medical Center (protocol 2010-P-000254/0) and Dana-Farber/Harvard Cancer Center (protocols 15-008 and 15-492). All patients provided informed consent before participating. This research was conducted in accordance with the principles of the Declaration of Helsinki.

### Histology

After resection, radical prostatectomy specimens were grossly examined and then formalin-fixed and paraffin-embedded (FFPE) according to standard procedures. Five-micron serial sections of tissue were mounted onto charged slides for staining or used directly for recovery of RNA. FFPE slides were stained with hematoxylin and eosin (H&E) using standard protocols. Quantification of residual tumor from treated cases was as previously reported [10, 11].

For IHC with antibodies against androgen receptor (AR), prostate-specific antigen (PSA), synaptophysin (SYP), NKX 3.1 and PSMA, slides were baked for 30□min at 60□°C, deparaffinized through xylenes and rehydrated through graded alcohols. Antigen retrieval was performed using a NxGen Decloaker (Biocare Medical), for 20□min at 95□°C in Tris-EDTA Buffer, pH 9.0 (Abcam; ab93684) for PSMA, PSA and SYP or for 15 min at 110□°C in Diva Decloaker (Biocare Medical; DV2004MX) for AR and NKX 3.1. After washing, a PAP pen border was drawn around tissue, and the sections were loaded into an intelliPATH FLX autostainer (Biocare Medical). Blocking was performed with 3% hydrogen peroxide (Fisher Scientific; BP2633) for 5 minutes and Background Punisher (Biocare Medical; BP974) for 10 minutes. Sections were then incubated with the primary antibody for 30 minutes at room temperature (PSMA: Dako M3620, diluted 1:500 into Renoir Red, Biocare Medical; PD904; NKX 3.1: Biocare Medical PP422AA, ready-to-use (RTU); AR clone D6F11: Cell Signaling 5153, diluted 1:200 into Renoir Red; PSA: Dako IR514, RTU; SYP: Dako M7315, diluted 1:200 into Renoir Red). Secondary labeling was performed with the Mach 4 Universal HRP Polymer Kit (Biocare Medical; M4U534). Colorimetric detection was achieved using Betazoid DAB (Biocare Medical; BDB2004) for 3 minutes, and counterstaining was performed using CAT Hematoxylin (Biocare Medical; CATHE). After dehydration through graded alcohols and clearing in xylenes, slides were mounted with Permount (Fisher Scientific, SP15).

After mounted slides were fully dried, residual mounting media was removed using xylene. Slides were then digitized on a Carl Zeiss AxioScan.Z1 microscope slide scanner equipped with a Plan-Apochromat 20× NA 0.8 objective, 266% LED intensity, 200 μs exposure time. Tissue images were acquired using ZEN Blue 2012 (Zeiss) with objective/magnification and pixel:distance calibrations recorded within the scanned CZI file.

### Manual image analysis

A 3 mm × 3 mm grid was overlaid onto each tissue section to divide the tissue into square regions of interest (ROIs) for analysis. Gridding was performed using the HALO Image Analysis Platform (Indica Labs). Each region was classified by PSMA staining and tumor presence in the area. Any region where tissue covered less than half of the area and contained no tumor or anti-PSMA positivity was given a class of 0 (blank). If most of the tumor within an ROI was positive for PSMA and most of the positivity was tumor, the region was given a class of 1 (true positive). If most of the tumor within an ROI was negative for PSMA, or if there was more negatively-stained tumor than positively-stained non-tumor, the region was classified as 2 (false negative). If most of the staining positivity in a region was non-tumor, the region was classified as 3 (false positive). If there was no tumor or staining positivity within a region, it was classified as 4 (true negative). Presence of any tumor or PSMA staining positivity not characterized by the primary score of each region was ignored. Classification of ROIs were verified by a board-certified and expert prostate pathologist (R.T.L.).

### Automated image analysis

The HALO Image Analysis Platform was used to quantify anti-PSMA stain intensity and area on a per-cell basis. From the treated cohort, ten PSMA-positive tumor ROIs and ten PSMA-positive non-tumor ROIs were randomly selected. Randomization was done to avoid multiple ROIs from the same patient selected for the same classification to prevent overrepresentation. ROIs were annotated to exclude non-type positivity (non-tumor staining excluded from an ROI representing tumor positivity) and intra-lumen secretion positivity. Indica Labs Area Quantification v2.1.11 analysis was then used to quantify the staining intensity and area of positivity. Analysis Parameters: stain 1 Color: 0.377, 0.424, 0.443; stain 2 Color: 0.164, 0.166, 0.121; stain 1 minimum optical density (OD): 0.2274, 0.427, 0.6708; blur radius: 6; minimum tissue OD 0.037. After analysis, the OD, relative areas of each stain’s intensity, and the overall staining area was recorded for each slide.

The Definiens Image Analysis Platform was used previously to quantify anti-AR and anti-PSA immunohistochemistry in post-treatment prostatectomy tissues [10]. This analysis produced IHC weighted intensity scores, or histology indices, based on pre-defined intensity thresholds on a per-cell basis. Each case intensity score is based on the formula:

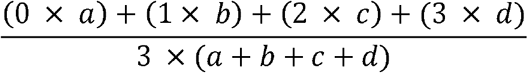

where *a* is the number of unstained cells, *b* is the number of weakly-stained cells, *c* is the number of moderately-stained cells, and *d* is the number of strongly-stained cells.

### Whole transcriptome sequencing analysis

Whole section ribbons of FFPE tissues were lysed in 1.5 ml microcentrifuge tubes using the RNeasy FFPE Mini Kit (Qiagen) according to the manufacturer’s instructions. 100-600 ng of recovered RNA was assembled into strand-specific, paired-end, Illumina-compatible sequencing libraries using the NEBNext Ultra II Directional RNA Library Prep Kit (New England Biolabs) with the NEBNext rRNA Depletion Kit (New England Biolabs). Libraries were quantified, pooled, and sequenced paired-end on a NovaSeq S4 flowcell with 100 cycles paired-end (2×100). Demultiplexed FASTQ files were adaptor-trimmed using Trimmomatic version 0.36 [12] and aligned to GRCh37 using RSEM version 1.3.2 [13] as a wrapper around STAR 2.7.0f [14], with an average of 24.9 million mapped reads (range: 8.0 to 47.8 million mapped reads) per sample.

Bulk RNA-seq counts were deconvolved to luminal cell-type specific values using the dampened weighted least squares (DWLS) method [15]. First, a cell signature matrix was generated using the *buildSignatureMatrixMAST* function using the single-cell RNA-seq dataset of human untreated prostate tumors by Karthaus *et al*. [16]. Annotation of cell types was performed by filtering by the percentage of mitochondrial genes, the number of unique molecular identifiers (UMIs), and the total number of UMIs. Potential doublets were removed. The resulting count matrix was normalized using *computeSumFactors* function from Scran version 1.2 [17], and integrated using the mutual nearest neighbor method with *fastMNN* from Batchelor version 1.8 [18]. Cells were clustered using the top 2000 genes with the most variance, and a total of eight cell types were identified using the cell type markers described in the original publication [16]: basal, luminal, endothelial, hillock, leukocytes, mesenchymal, smooth, and T cell. To impute the luminal cell-type expression of *FOLH1*, we applied the formula:

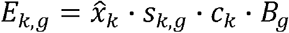

where *E*_*k,g*_ is the expression of gene *g* in counts, derived from cell type 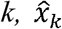 is the estimated percentage of cell type *k* in the bulk sample, *s*_*k,g*_ is the relative expression of gene *g* in cell type *k* derived from normalized, untransformed scRNA-seq, *c*_*k*_ is the cell size of cell type k, and *B*_*g*_ is the total gene count of *g* from the bulk RNA-seq. Deconvolved values were represented as counts per million (CPM). These analyses were performed using R version 4.0.5 and RStudio version 1.4717.

### Statistical analysis

Statistical analyses were performed with GraphPad Prism version 9 (GraphPad Software) for Mac. Associations between factors were measured using Pearson or Spearman correlations. Comparisons of single factors between treated and untreated tumors, between regions of tumor and non-tumor, or between exceptional responder (ER) and incomplete and nonresponder (INR) cases were performed using Welch’s *t* tests. Null hypothesis tests of enrichments between treated or untreated tumors and individual dichotomous factors were performed using two-sided Fisher’s exact tests. Comparisons of covariates within single factors between treated and untreated tumors were performed using Cochran-Armitage tests for trend. The accuracy of PSMA immunohistochemistry for distinguishing INR from ER cases was examined using a receiver operating characteristic curve. Statistical significance was pre-specified at *P* < 0.05. Grid-based detection performance was reported for treated and untreated cases with 95% confidence intervals, obtained from the 2.5% and 97.5% percentiles from 2,000 bootstrap samples by a random sampling of patients with replacement. The significance of the difference in performance metrics between treated and untreated cases was calculated by Wald tests.

## RESULTS

### Immunohistochemistry against prostate-specific membrane antigen identifies residual tumor following neoadjuvant intense androgen deprivation therapy

We assessed the presence of residual tumor in prostatectomy specimens following six months of systemic neoadjuvant intense androgen deprivation therapy (iADT) in a cohort of 35 patients as part of a phase 2 clinical trial [10]. Although hematoxylin and eosin (H&E) staining of whole-mount surgical specimens was generally sufficient to detect most of the residual adenocarcinoma in patients, immunohistochemistry (IHC) was employed for confirmation of residual disease, detection of individual tumor cells, and precise calculations of post-treatment tumor volumes. Staining for the common prostate cancer markers PSA and AR displayed reduced staining in a subset of cases due to suppression of AR by iADT (Fig. 1A-B) and thus was not as useful in this post-treatment setting. In some cases, diffuse staining of the linage marker NKX 3.1 helped to identify residual tumor (Fig. 1A). In almost every case, however, immunostaining with antibodies against PSMA demonstrated strong and consistent staining of residual tumor (Fig. 1A-B). In these post-treatment tumors, anti-PSMA IHC was weak in benign glands (Fig. 1C). By contrast, prostatectomy specimens from patients not treated with androgen deprivation therapy displayed stronger PSMA staining in both benign and tumor glands (Fig. 1D).

**Figure 1.**
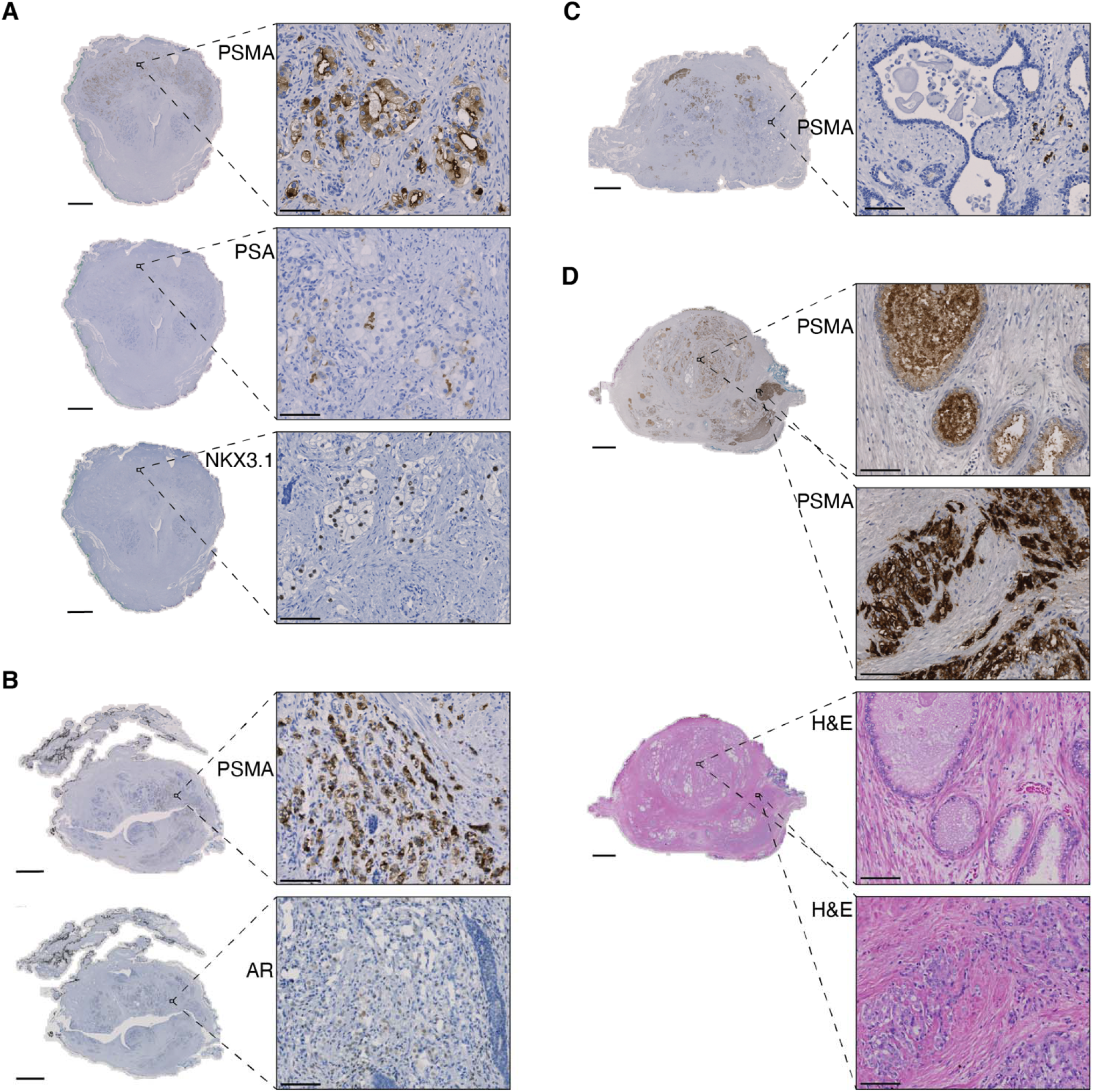
Representative whole-slide and inset micrographs of prostate tumor. (A) Residual tumor positive for anti-PSMA, negative for anti-PSA, and positive for anti-NKX 3.1. (B) Residual tumor positive for anti-PSMA and diffusely weak for anti-AR. (C) A region of benign glands shown are negative for anti-PSMA adjacent to PSMA-positive residual tumor cells. (D) Untreated prostatectomy tissue with both benign (top) and tumor (bottom) glands staining positively for anti-PSMA. H&E is shown for reference. Whole-slide image bar: 5 mm; inset bar: 100 μm.

Given that radiolabeled ligands for PSMA protein are now available for positron emission tomography (PET) for tumor localization *in vivo*, we asked whether our detection of residual tumor via IHC would mimic potential detection by PSMA-PET. Downsampling each case into a 3 mm × 3 mm grid to approximate the resolution of PET, we devised a straightforward scoring scheme to test the cohort systematically. These 9 mm^2^ regions of interest (ROIs) containing tumor would either be marked as positively stained by PSMA (true-positive) or not stained (false-negative) (Fig. 2A). Similarly, regions without tumor could be stained by PSMA (false-positive) or not stained (true-negative). An example of this gridding scheme is shown in Figure 2B. From our cohort of 37 patients that completed treatment, one prostatectomy specimen encountered technical delays in sample processing and was not suitable for immunohistochemical analysis. A second patient underwent TURP instead of prostatectomy, leaving 35 treated prostatectomy specimens for analysis. As controls, we assembled a cohort of 37 patients comprised of intermediate-to-high risk prostate cancer treated only by surgery (Table 1). However, since iADT trials select for higher risk patients than are normally treated by surgery alone, higher Gleason Grade Groups in the treated cohort were unavoidable (Table 1). Representative ROIs for each of the possible scores are shown in Figure 2C for the treated cohort and Figure 2D for the untreated cohort.

**Figure 2.**
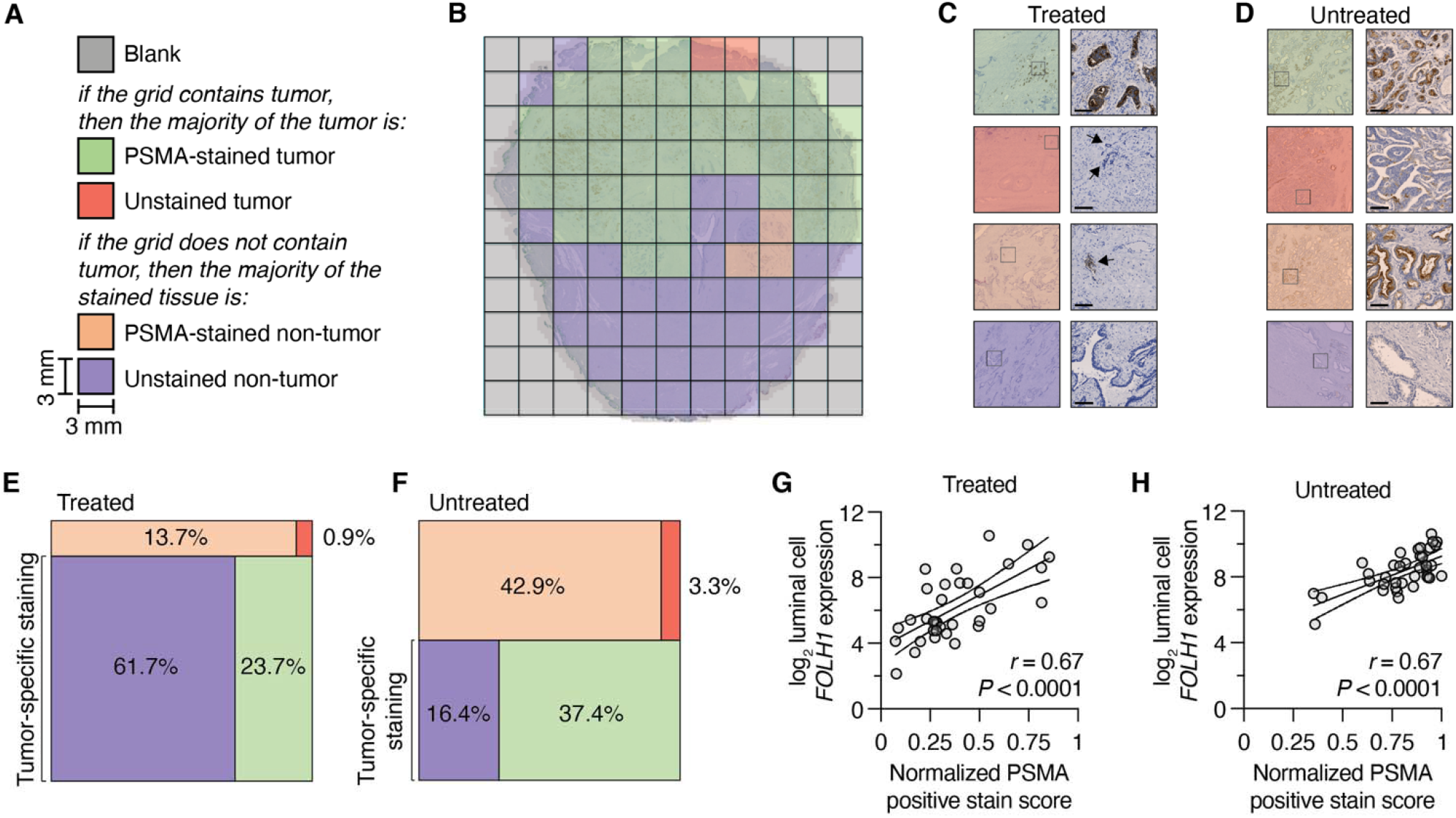
Scoring of prostate tumor tissues for PSMA staining. (A) Color coding of regions of interest (ROI) into 9 mm^2^ grids. (B) Example of a whole mount treated prostate tissue gridded and scored. (C-D) Representative ROIs from a treated (C) or untreated (D) prostate tumor. Inset bar: 100 μm. (E-F) Visualization of distribution of ROIs across the treated (E) or untreated (F) cohort, normalized per-patient. (G-H) Pearson correlation of the proportion of positive-staining regions (tumor and nontumor) per-patient with the deconvolved luminal cell expression of *FOLH1* in counts per million (CPM) from bulk RNA sequencing of a serial slide of the same tissue block from the treated (G) or untreated (H) cohort. Lines indicate linear regression trend with 95% confidence interval.

**Table 1.** Patient characteristics.

|  | *Treated* | *Untreated* | *P value* |
| --- | --- | --- | --- |
| *N* | 35 | 37 |  |
| Age (years), median (IQR) | 62 (58-69) | 63 (58-66) | 0.81<sup>a</sup> |
| Race, *n* (%) |  |  |  |
| White | 24 (69%) | 28 (76%) | 0.55<sup>b</sup> |
| Black | 5 (14%) | 5 (14%) |  |
| Asian | 2 (6%) | 0 (0%) |  |
| Hawaiian/Pacific Islander | 0 (0%) | 1 (3%) |  |
| Unknown | 4 (11%) | 3 (8%) |  |
| Baseline PSA (ng/ml), median (IQR) | 9.31 (5.53-17.29) | 6.18 (4.65-7.54) | 0.19<sup>a</sup> |
| Final pathologic yT/T stage, *n* (%) |  |  |  |
| T0 | 4 (11%) | N/A (N/A) | 0.25<sup>b</sup> |
| T2 | 18 (51%) | 15 (41%) |  |
| T3a | 5 (14%) | 16 (43%) |  |
| T3b | 7 (20%) | 6 (16%) |  |
| T4 | 1 (3%) | 0 (0%) |  |
| N1 | 5 (20%) | 3 (8%) | 0.47<sup>c</sup> |
| ISUP Grade Group (biopsy/final pathology), *n* (%) |  |  |  |
| 2 | 4 (11%) | 5 (14%) | < 0.001<sup>b</sup> |
| 3 | 4 (11%) | 26 (70%) |  |
| 4 | 11(31%) | 3 (8%) |  |
| 5 | 16 (46%) | 3 (8%) |  |
| Molecular alteration by immunohistochemistry, *n* (%) |  |  |  |
| PTEN loss/reduction | 20 (57%) | 14 (38%) | 0.16<sup>c</sup> |
| Nuclear ERG positive | 14 (40%) | 13 (35%) | 0.81<sup>c</sup> |
| Tumor content of slide (%), median (IQR) | 10 (2-20) | 25 (17.5-47.5) | < 0.001<sup>a</sup> |

<sup>a</sup> Welch's *t*-test.<sup>b</sup> Cochran-Armitage test for trend.<sup>c</sup> Fisher's exact test.

One of the major differences between the treated and untreated cohort was the volume of tumor per slide (Table 1), with the treated cohort harboring substantially fewer ROIs containing tumor (24.6% *vs*. 40.7%; Fig. 2E-F). The majority of ROIs in the treated cohort was PSMA-unstained non-tumor (Fig. 2E) whereas the largest proportion of ROIs in the untreated cohort was PSMA-stained non-tumor (Fig. 2F). In the treated cohort, 85.0% of the ROIs demonstrated PSMA staining specific for tumor (Fig. 2E) vs. 53.9% in the untreated cohort (Fig. 2F), which we utilized for the purposes of identifying tumor only (Table 2). While sensitivity, specificity, positive predictive value (PPV) and negative predictive value (NPV) were also all greater for treated tumors (Table 2), only accuracy, specificity and NPV differences were statistically significant. The total positively-stained fraction (encompassing both true- and false-positives) was 37.4% for the treated cohort and 80.3% for the untreated cohort.

**Table 2.** Grid-based ROI anti-PSMA performance summary. 95% confidence intervals calculated from 2000 bootstrap samples (with replacement) at the patient level. *P* values determined by Wald tests.

|  | *Treated* | *Untreated* | *P value* |
| --- | --- | --- | --- |
| **Accuracy** | 0.850 (0.805-0.891) | 0.539 (0.466-0.607) | < 0.0001 |
| **Sensitivity** | 0.955 (0.911-0.986) | 0.914 (0.840-0.968) | 0.30 |
| **Specificity** | 0.816 (0.763-0.866) | 0.278 (0.216-0.351) | < 0.0001 |
| **Positive Predictive Value** | 0.629 (0.477-0.759) | 0.468 (0.378-0.550) | 0.06 |
| **Negative Predictive Value** | 0.982 (0.962-0.995) | 0.824 (0.727-0.918) | 0.0012 |

With the marked differences between cohorts for the extent of positive tumor staining, we therefore sought to validate this finding orthogonally. From a serial slide from each block, we collected the entire tissue (encompassing both benign and tumor glands, as well as stromal cells) and performed whole transcriptome sequencing. RNA-seq was performed for each case in both cohorts. These bulk transcriptomes were then deconvolved to obtain luminal epithelial cell-specific gene estimates without distinguishing tumor from benign cell types. On a per-sample basis we then correlated the anti-PSMA positive stain score with the corresponding expression value for *FOLH1* (*folate hydrolase 1*), the gene encoding PSMA. Strikingly, these associations were very strongly positive in both cohorts (Fig. 2G-H), with a Pearson coefficient of correlation (*r*) of 0.67 (95% C.I. 0.43-0.82) for the treated cohort and 0.67 (95% C.I. 0.44-0.81) for the untreated cohort. Thus, our histologic and low-resolution estimates of PSMA protein expression in tissue were proportional to values derived from mRNA expression and thus validate the accuracy of using IHC to assess epithelial cell expression of PSMA.

### Factors influencing analysis

The treated cohort invariably contained less tumor due to the effect of treatment, and thus the fraction of ROIs containing any tumor were significantly less than the untreated cohort (Fig. 3A, *P* = 0.0061, Welch’s *t* test). Across both cohorts, the rates of accuracy and PPV increased proportional to the amount of tumor on the slide (Fig. 3B), with untreated tumors showing stronger proportional relationships by Pearson correlation (*r* = 0.80 and *r* = 0.92, respectively). However, even in treated cases with very low tumor volumes, the accuracy rate remained high (Fig. 3B) due to the lower false-positive rate of benign glands demonstrating positive staining for PSMA. We then compared the relative staining intensity between PSMA-positive benign and tumor cells. We employed the HALO image analysis platform on ten randomly selected ROIs of tumor and benign glands from ten different cases and developed a cell analysis algorithm to quantify staining (Fig. 3C). The intensity of staining was greater in tumor versus benign, as was the optical density (Fig. 3D). Within each 9 mm^2^ ROI, individual PSMA-stained tumor cells had a greater stained area than PSMA-stained non-tumor cells (Fig. 3D). Thus, when accounting for the likelihood of greater numbers of positive tumor cells per 9 mm^2^ ROI than positive non-tumor cells, our downsampled analysis was more likely to underestimate the accuracy and specificity relative to an exact per-cell annotation.

**Figure 3.**
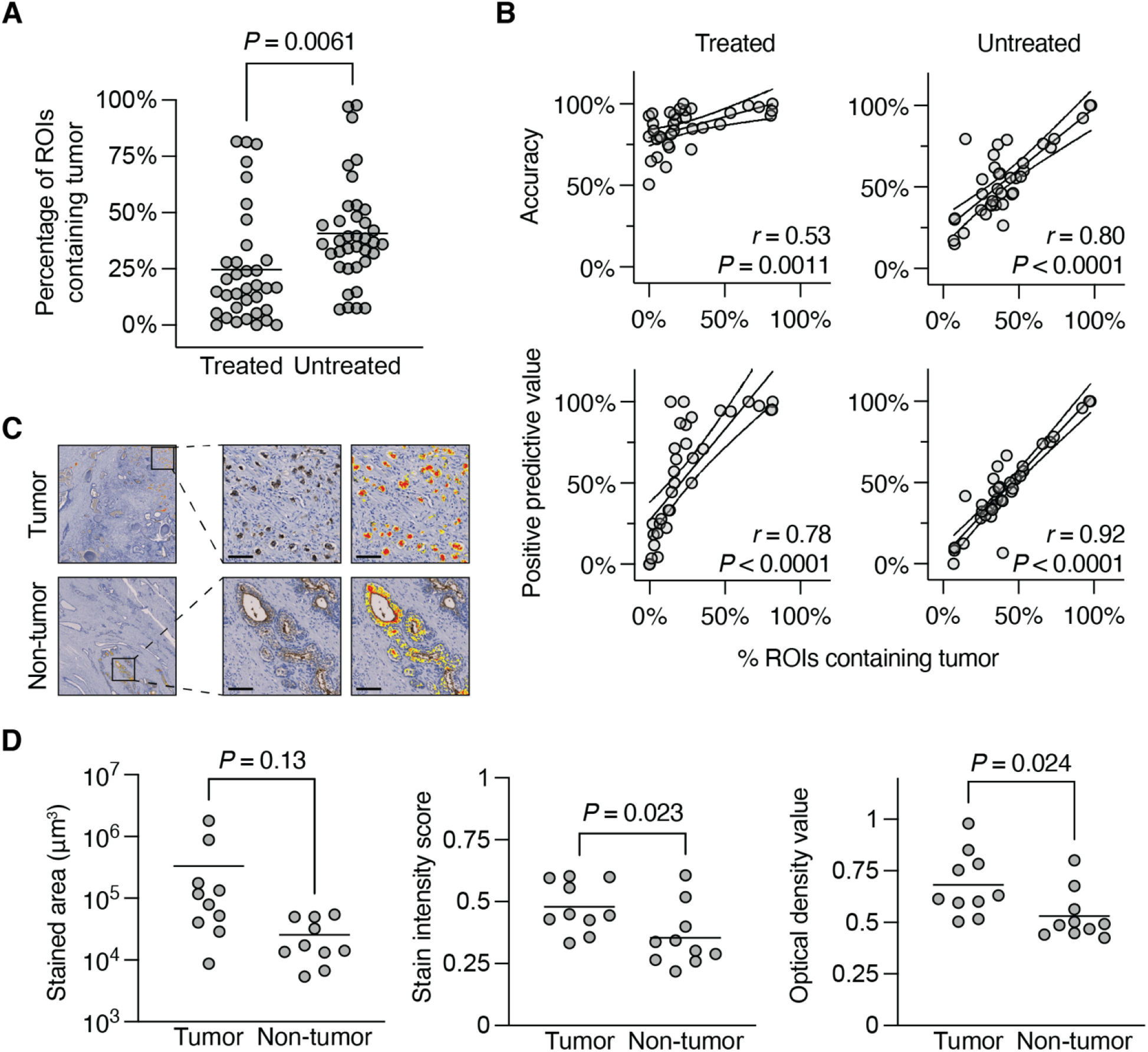
Assessment of factors influencing accuracy of anti-PSMA to detect tumor cells. (A) Dot plot comparing the proportion of ROIs containing tumor between the treated and untreated cohorts by Welch’s *t* test. Line indicates mean. (B) Pearson correlations of the accuracy of anti-PSMA to only identify tumor cells (top) and the positive predictive value of anti-PSMA to identify tumor cells (bottom) as a function of the proportion of ROIs containing tumor. Lines indicate linear regression trend with 95% confidence interval. (C) Representative ROIs of tumor (top) or non-tumor (bottom) selected for per-cell analyses using HALO. Inset bar: 100 μm. (D) Dot plots comparing the stained area (left), staining intensity score (middle) and optical densities (right) from each HALO per-cell analysis, by Welch’s *t* test. Line indicates mean.

### Benefits of anti-PSMA immunohistochemistry for detecting residual tumor

Robust and thorough identification of residual disease is critical for accurate quantification of post-treatment tumor volumes. Although large tumor foci were seldom challenging to quantify, rare and scattered single cells present significant visual challenges using H&E and conventional stains. Minimal residual disease typically is measured in one dimension, defined as from 0.25 to 0.5 cm. In our clinical trial cohort, incomplete or nonresponding patients (INR) were defined as those patients with tumor volumes greater than 0.05 cm^3^. These cases often displayed scattered single cells with the same relative frequency as exceptional responders (ER, residual tumor volumes less than 0.05 cm^3^). These scattered cells were readily positively-stained by anti-PSMA IHC (Fig. 4A) and were distinguishable from benign glands and stroma by histology as well. We next considered whether total PSMA staining of any cell type would distinguish ER from INR patients. The combined PSMA positivity of tumor and non-tumor was greater in INR than ER (Fig. 4B, *P* = 0.0084, Welch’s *t* test) and identified cases with greater residual tumor volumes with an area under the receiver operating characteristic curve (AUC) of 0.72 (Fig. 4C).

**Figure 4.**
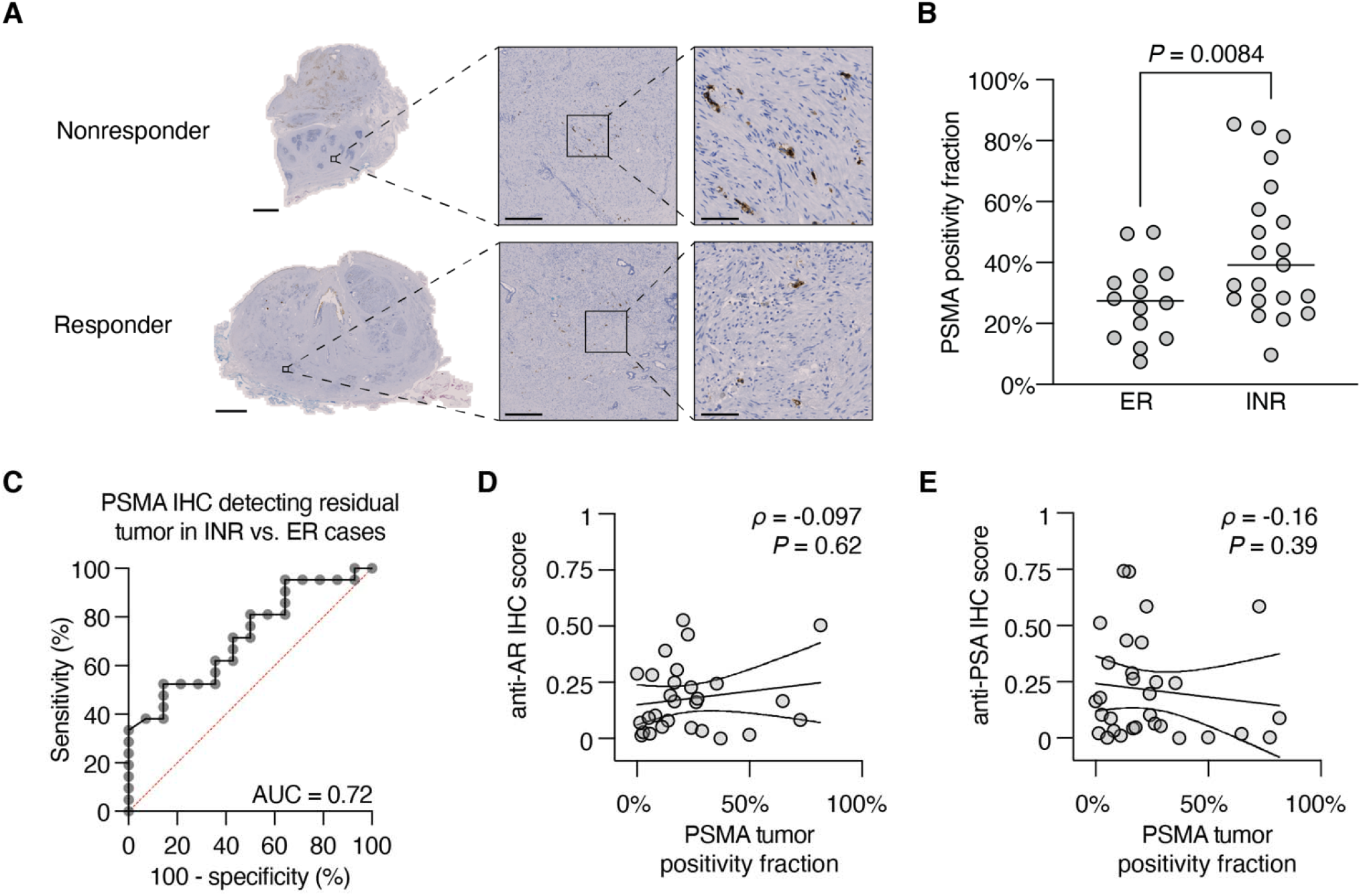
Benefits of anti-PSMA immunohistochemistry for detecting residual tumor. (A) Representative micrographs of a nonresponder and responder case showing anti-PSMA detection of rare scattered residual tumor cells. Whole-slide image bar: 5 mm; low-power inset bar: 500 μm; high-power inset bar: 100 μm. (B) The total PSMA positivity fraction (ROIs containing PSMA-positive benign or tumor) comparing exceptional responders (ER) and incomplete and nonresponders (INR) by Welch’s *t* test. Line indicates mean. (C) Receiver operating characteristic curve for the ability of the PSMA positivity fraction to distinguish ER from INR cases. (D) Spearman correlation of tumor-specific anti-AR nuclear intensity index (D) or anti-PSA cytoplasmic intensity index (E) *vs*. the fraction of ROIs with tumor-specific anti-PSMA positivity. Lines indicate linear regression trend with 95% confidence interval.

As iADT functions to suppress AR expression and its activity, we previously assessed the expression of AR and its target gene product PSA by IHC on the Definiens quantification platform as a direct phenotypic outcome of treatment [10]. Relative to untreated control biopsy tissues, both anti-AR and anti-PSA displayed significant reductions in histology intensity index scores in treated tumors [10], with the vast majority of cases showing low to no expression. Not surprisingly, these intensity scores showed no significant association with PSMA tumor-specific positivity by Spearman correlation (Fig. 4D-E).

Finally, we asked whether anti-PSMA IHC had the capability to detect rarer treatment-induced phenotypes involving neuroendocrine differentiation in the treated cohort. Amongst the subtypes of prostate cancer with neuroendocrine differentiation proposed by the Prostate Cancer Foundation working committee and 2016 World Health Organization classifications [19], we identified two cases with foci of neuroendocrine differentiation in our cohort: adenocarcinoma with Paneth cell-like neuroendocrine differentiation (Fig. 5A) and high-grade neuroendocrine carcinoma with small cell carcinoma-like features (Fig. 5B). These foci in both of these cases displayed strong positive staining for the neuroendocrine marker synaptophysin (SYP) and were negative for AR. Remarkably, both retained anti-PSMA expression, albeit with more diffuse staining patterns in these higher-grade components. Our cohort also included a single case harboring amphicrine carcinoma [20], which retained positivity for anti-AR IHC and also displayed robust anti-PSMA positivity (Fig. 5C). Thus, anti-PSMA IHC was able to accurately detect these poorly differentiated foci of treatment resistant tumor.

**Figure 5.**
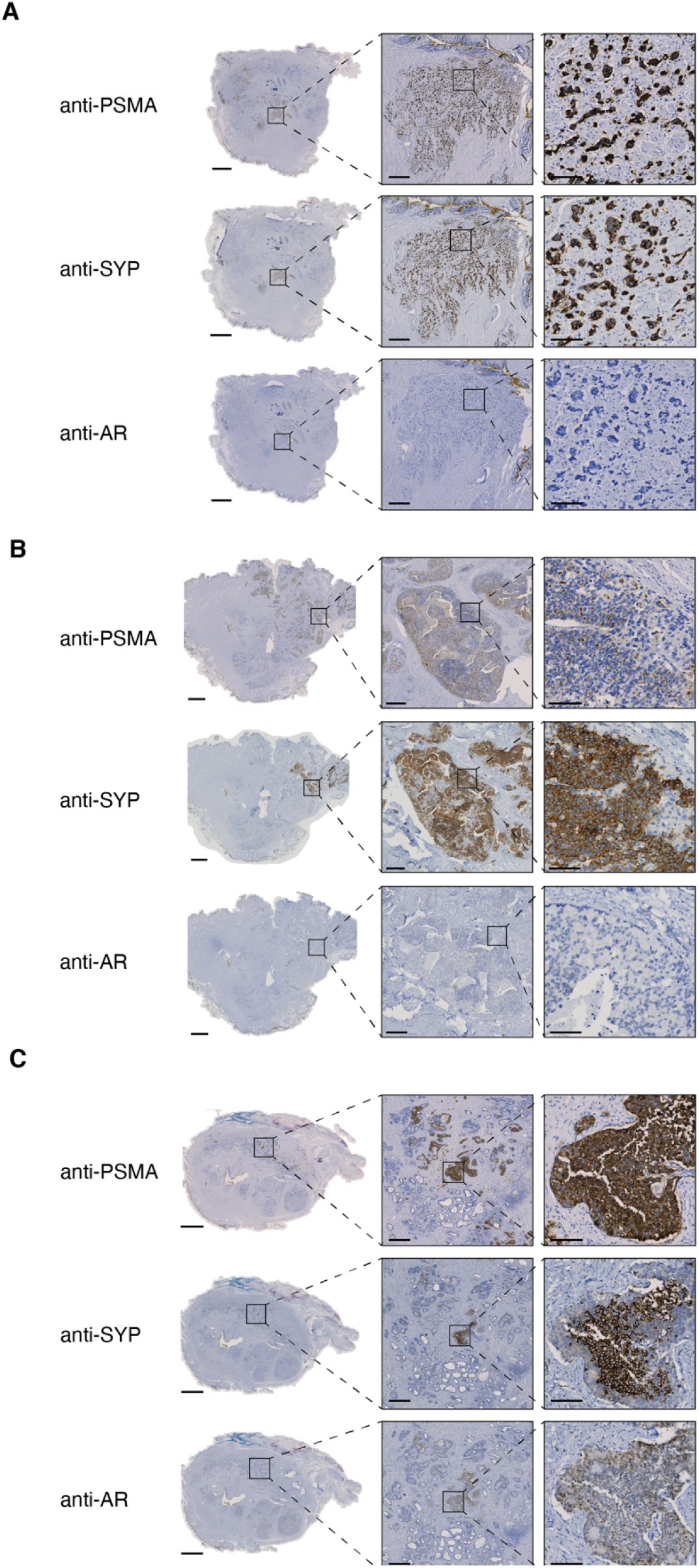
Persistent PSMA expression in residual tumor with neuroendocrine features. anti-PSMA, anti-Synaptophysin (SYP), and anti-AR immunohistochemistry in residual tumors harboring (A) Paneth cell-like morphology, (B) high-grade small-cell neuroendocrine carcinoma, or (C) amphicrine carcinoma. Whole-slide image bar: 5 mm; low-power inset bar: 500 μm; high-power inset bar: 100 μm.

## DISCUSSION

Intense androgen deprivation can increase the expression of other genes in the context of diminished AR activity. By contrast, the upregulation of prostate-specific membrane antigen (PSMA) (protein product of the *FOLH1* gene) is a well-established prostate tumor-specific response to ADT [21]. *FOLH1* encodes a type II transmembrane glutamate carboxypeptidase with markedly strong expression in prostatic epithelial cells [22]. *FOLH1* is predominantly expressed in the prostate, with much lower levels of expression in the brain, salivary gland, and small intestine [23], as well as in the neovasculature of solid malignancies [24]. PSMA is expressed in benign prostatic epithelium, primary cancer, and most lymph node metastases [23], with higher expression in primary prostate tumors and metastatic lesions compared to benign tissue [25]. Unlike the AR-responsive *KLK3*, however, *FOLH1* increases in expression inversely proportional to AR activity [21].

The identification of this gene’s upregulation in advanced prostate cancers has made it is an ideal target for both molecular imaging and precision molecular radiotherapy [26]. However, its utility as a marker for treatment response after neoadjuvant therapy with second-generation AR-axis inhibitors has not been explored. In this study, we quantified the extent of tumor-specific anti-PSMA immunoreactivity in the context of downsampling a 20× whole-scanned slide to 3 mm × 3 mm regions of interest (ROIs) in prostate sections following six months of ADT plus enzalutamide. We found that anti-PSMA IHC was highly specific for residual tumor but not benign glands, and that total anti-PSMA reactivity was highly correlated with expression of *FOLH1*. In addition, anti-PSMA IHC identified regions of tumor that were negative for other known markers of prostate including AR and PSA, and also marked regions of synaptophysin-positive neuroendocrine differentiation.

For patients receiving iADT, inhibition of AR activity results in downregulation of AR-regulated genes including *KLK3*, which encodes PSA. Antibodies against PSA are usually sensitive for detecting rare prostate cancer cells in prostatectomy tissue but anti-PSA sensitivity suffers after inhibition of AR and thus can be suboptimal for identifying rare scattered cells with treatment effect [27]. Lineage markers such as NKX 3.1 are similarly useful in the untreated setting, but potential deletions to *NKX3-1* on chromosome 8p selected by therapy can limit its utility as a marker of residual disease [28, 29]. Although decreased AR activity rarely decreases the expression of basal (p63) or luminal (high molecular weight cytokeratin) markers, the expression of these markers is not specific to benign cells, and tangential sectioning of atrophic benign glands can be a mimicker for residual tumor [30, 31].

The purpose of downsampling was to approximate the resolution of PET imaging using either ^18^F-DFCPyL or ^68^Ga-PSMA-11, two PSMA-PET tracers approved in 2021 by the United States Food and Drug Administration for *in vivo* localization of prostate cancer in patients with a high risk of metastasis [32, 33]. The approval of these agents is based on the superior sensitivity of PSMA-PET to detect prostate tumors in both newly-diagnosed localized and biochemically recurrent settings, even amongst lesions not detected by multiparametric MRI [34, 35]. Consistent with these previous reports, we observed greater PSMA intensity in regions of tumor compared to nontumor in both cohorts, but absolute PSMA reactivity was non-existent in most non-tumor ROIs in the treated cohort only, markedly increasing the specificity PSMA after treatment (see Table 2). This finding highlights the potential utility of PSMA imaging for monitoring *in vivo* treatment response during iADT neoadjuvant therapy. In particular, our observation of distinctive retention or up-regulation of PSMA in tumor cells with its down-regulation in benign glands is supportive of at least two ongoing trials of neoadjuvant iADT with on-study PSMA scans (NCT03860987 and NCT03080116) and the added benefit of sectioning whole-mount prostatectomy specimens in the same plane as the PET/MR.

Of particular note, we observed specific PSMA expression in three separate cases harboring residual cancer with evidence of neuroendocrine differentiation. In two of these cases, reconstitution of AR activity was expected with either Paneth cell-like change or amphicrine features, retaining most exocrine markers [19]. However, the detection of PSMA by IHC, albeit reduced, in a residual tumor exhibiting high-grade neuroendocrine small-cell morphology was unexpected. Indeed, PSMA expression was reduced in treatment-induced neuroendocrine prostate cancers compared to castration-resistant prostate adenocarcinomas in several cohorts [36-38], and androgen deprivation of prostate adenocarcinoma cell lines recapitulated this effect *in vitro* [39]. In our treated cohort case exhibiting high-grade neuroendocrine small-cell morphology, synaptophysin expression was focal across the remainder of the tissue (see Fig. 5B, whole-slide), with reduced or diffuse PSMA expression throughout all foci of residual tumor. This pattern suggests that these SYP-positive foci are still in transition with six months of treatment, and do not represent a fully-differentiated neuroendocrine tumor. In contrast to castration-resistant prostate cancer, where prolonged ADT treatment can select for PSMA-negative, SYP-positive neuroendocrine metastases that are invisible to PSMA-PET, the shorter duration of neoadjuvant intense ADT indicates that PSMA-PET has the potential to detect all residual tumors.

In light of our findings, we wish to highlight important limitations of this study. First, the untreated cohort was not comprised of matched biopsies acquired prior to treatment from the same patients as the treated cohort. Although we may have readily observed similar results from those tissues rather than using a separate cohort, our clinical study’s targeted biopsy approach would not have sampled sufficient numbers of benign glands to make an adequate and robust comparison. We thus strove to compare prostatectomies to prostatectomies. The second limitation is that prostate tissues from the untreated cohort harbored a greater proportion of tumor glands per slide (see Fig. 2), but our analyses were adjusted to account for patient-level effects on tumor content. The amount of tumor per slide did influence the unadjusted analysis, but the inherent nature of a comparable untreated cohort to our treated group would mean that we would have needed to identify a greater proportion of lower-risk tumors to match tumor volumes between groups. iADT trials select for higher-risk patients, so by default the treated cohort harbored higher Gleason Grade Groups despite comparable pT stages (see Table 1). Finally, neither cohort underwent PSMA-PET imaging (DCFBC or DCFPyL), so actual correlation with *in vivo* imaging was not possible, and frozen whole-mount specimens were not available for possible correlation with PET tracer binding by autoradiography.

As more clinical studies use pathologic responses as primary endpoints, more accurate means to measure volumes of residual tumor become paramount. For neoadjuvant iADT trials, these tumor volumes distinguish responder from nonresponder status, may predict disease recurrence risk, and may be used for adjuvant therapy stratification in the future. In the current study, we showed that anti-PSMA IHC alone had 96.6% sensitivity and 81.8% specificity for detecting residual disease when each ROI was separately examined by an expert prostate pathologist using H&E and other confirmatory stains. By contrast we previously showed that iADT reduces expression of PSA by IHC by approximately 3-fold such that approximately 35% of cases do not express detectable levels of PSA *in situ*, with similar observations for anti-AR IHC, relative to matched untreated controls [10]. These findings are largely in agreement with prior reports of short- and long-course neoadjuvant hormone therapy for prostate cancer [3, 27, 40]. Thus, we propose that anti-PSMA should be considered as a standard immunohistochemical marker for identifying difficult-to-visualize residual tumors, for measuring residual tumor volumes, and assessing overall pathologic response to neoadjuvant iADT.

Significantly, there is an unmet opportunity for artificial-intelligence (AI)-driven efforts to use anti-PSMA IHC for refining algorithms currently used for detecting suspicious regions in untreated prostate tumors [41-43]. Accurate reporting of neoadjuvant hormone-treated tumors has remained a challenge for pathologists for years, and current diagnostic efforts continue to be plagued by atypical histologies requiring further expert pathologic review, with these tumors more likely to be missed by automated systems or without specialized immunostains [27, 44]. Larger phase 3 studies of neoadjuvant iADT (*i*.*e*. NCT03767244) may soon delineate progression-free benefits to neoadjuvant therapy with or without adjuvant hormonal treatment, which may be stratified by residual cancer burdens. We have shown here that anti-PSMA is a reliable marker for post-treatment prostate tumors, and broader utilization of this stain may identify patients who would benefit from intensified adjuvant therapies by more accurately documenting their residual tumor burden.

## Data Availability

The data underlying this article are available in the Database of Genotypes and Phenotypes (dbGaP) and Gene Expression Omnibus (GEO) at https://www.ncbi.nlm.nih.gov/gap/ and https://www.ncbi.nlm.nih.gov/, respectively, and can be accessed with phs001813.v2.p1 (dbGaP), phs001938.v3.p1 (dbGaP), GSE183100 (GEO) and GSE183019 (GEO).

https://www.ncbi.nlm.nih.gov/geo/

## ACKNOWLEDGMENTS

The authors gratefully acknowledge the patients and the families of patients who contributed to this study. The authors thank Dr. Margaret White for her helpful discussions and critical review of the manuscript. Portions of this work utilized the computational resources of the NIH HPC Biowulf cluster.

## Notes

**COMPETING INTERESTS** R.T.L. performs consulting in an advisory role for Janssen Pharmaceuticals. The remaining authors declare no conflicts of interest.

**FUNDING** This work was supported by the Prostate Cancer Foundation (Young Investigator Award to S.W.), the Department of Defense Prostate Cancer Research Program (W81XWH-19-1-0712 to S.W., W81XWH-16-1-0433 to A.G.S.), and the Intramural Research Program of the NIH, National Cancer Institute.

### Competing Interest Statement

R.T.L. performs consulting in an advisory role for Janssen Pharmaceuticals. The remaining authors declare no conflicts of interest.

### Clinical Trial

NCT02430480

### Funding Statement

This work was supported by the Prostate Cancer Foundation (Young Investigator Award to S.W.), the Department of Defense Prostate Cancer Research Program (W81XWH-19-1-0712 to S.W., W81XWH-16-1-0433 to A.G.S.), and the Intramural Research Program of the NIH, National Cancer Institute.

### Author Declarations

The collection and analysis of tissue and demographic data from patients with high-risk localized prostate cancer treated with ADT plus enzalutamide prior to surgery was approved by the National Institutes of Health Institutional Review Board (protocol 15-c-0124). The collection and analysis of tissue and demographic data from patients with localized prostate cancer treated only by surgery was approved by the institutional review boards of Beth Israel Deaconess Medical Center (protocol 2010-P-000254/0) and Dana-Farber/Harvard Cancer Center (protocols 15-008 and 15-492).

